# Timely Identification of deteriorating Patients from acute respiratory infections at the primary care level in the COVID-19 Era: quality improvement collaborative

**DOI:** 10.1101/2025.02.12.25321948

**Authors:** Facundo Jorro-Barón, Andrea Falaschi, Lía Bosio, Luz Gibbons, Emilse Vitar, Marina Guglielmino, Erica Negri, Belén Peralta-Roca, Ana Paula Rodriguez, Inés Suarez-Anzorena, Juan Pedro Alonso, Viviana Rodriguez, Javier Roberti, ICARO study group, Ezequiel García-Elorrio

**Author notes:** Corresponding author: Facundo Jorro-Baron MD, MsC(c), Researcher, Quality and Safety in Healthcare, Institute for Clinical Effectiveness and Health Policy, 2024 Ravignani, (1414) Buenos Aires Argentina. PFIZER COMPETITIVE GRANT PROGRAM ID: 74261789.

## Abstract

**Background:** Primary care has been essential in ensuring the continuity of health services for patients with COVID-19 and other conditions. We aimed to increase the adoption of evidence-based interventions to identify clinical deterioration in adult patients with confirmed or suspected respiratory COVID-19 at the primary care level.

**Methods:** We implemented specific interventions in nine Primary Health Care Centers (PHCC) through a Quality Improvement Collaborative (QIC) with an interrupted time-series design. Interventions included triage for acute respiratory symptoms, the NEWS2 scale, portable oximeters for selected patients, and the provincial telehealth system. Additional components involved leadership commitment, teamwork tools, reminders, audits, feedback, and direct observation. A mixed-method evaluation was conducted, with two learning sessions and three action periods to test and implement selected change ideas.

**Results:** Six PHHCs completed the study. Over 48 weeks, data from 877 patients were gathered, 356 in the baseline period (BP) and 477 in the implementation period (IP). Eight hundred sixty-two medical consultations were reported, 367 for BP and 495 for IP. More COVID-19-confirmed diagnoses were observed in the IP group (1.9% vs 15%, p<0.001).

The bundle was implemented in 0% and 28.4% of patients in the BP and IP groups, respectively. Upon evaluating the individual components of the bundle, we discovered enhancements in the utilisation of triage, application of NEWS2, and utilisation of oximeters when appropriate. A decrease in the number of follow-up calls was observed at the end of the implementation.

Patients rated the quality of care as positive in 66% of the cases in the BP and 76% in the IP group (p=0.023).

**Conclusion:** We successfully implemented a triage algorithm based on the NEWS2 score to identify respiratory deterioration in adult patients in primary care through a QIC. This intervention was perceived as an improvement in the quality of care by the patients.

**What is already known on this topic:** There is an urgent need to develop new methods to support patients with SARS-CoV-2 or other respiratory infections at risk of developing severe disease, implement new remote care models to facilitate risk stratification, decompress hospitals and emergency rooms, and preserve the availability of personal protective equipment.

**What this study adds:** Through a Quality Improvement Collaborative (QIC), we implemented a triage algorithm based on the NEWS2 score to identify respiratory deterioration in adult patients in primary care.

This intervention improved the patient’s perceptions of their quality of care.

**How this study might affect research, practice or policy:** The utilisation of the triage algorithm founded on NEWS2 in primary care settings has garnered extensive approval, thereby enabling the expansion of interventions to additional locations within the region.

## BACKGROUND

The SARS-COV-2 pandemic exposed a complex matrix in which demand exceeded the capacity of hospital systems and resources, particularly in low-income countries[1]. Coronavirus disease-19 (COVID-19) has a broad spectrum of severity; in most cases, the infection results in a self-limiting disease that can be managed on an outpatient basis without any specific medical intervention. However, the need for hospital treatment is associated with a significant increase in mortality[2]. The risk of adverse outcomes was not equally distributed across the study population. Argentina had more than 9 million confirmed cases with a mortality rate of 1.31%[3]. Because of this overwhelming situation, COVID-19 has required a large-scale reorganisation of existing acute care pathways.

Health systems have adopted different strategies to mitigate the negative impact of COVID-19. There is an urgent need to develop new methods to support patients with SARS-CoV-2 or other respiratory infections at risk of developing severe disease, implement new remote care models to facilitate risk stratification, decompress hospitals and emergency rooms, and preserve the availability of personal protective equipment [4]. These new models of care should have strict infection control measures, unlimited access to PPE, the ability to perform viral tests, and the incorporation of a degree of redundancy in workforce planning. Traditional face-to-face consultations have been replaced by remote triage whenever possible [5].

### RATIONALE

Regarding Argentina’s health system, hospitals receive and treat patients with health conditions that could be managed at the primary care (PC) level. Possible explanations for this situation could be the lack of trust in health providers from the users’ perspective due to the perceived lack of skills to manage different clinical conditions, the lack of confidence in the PC due to long waiting times, problems getting appointments, poor quality of care, and poor customer service[6].

Importantly, outpatient management is appropriate for most COVID-19 patients. In most cases, the disease is mild and does not require medical intervention and/or hospitalisation, particularly in vaccinated individuals. However, some patients may develop serious illnesses that require hospital treatment. Identifying patients who are likely to need hospital treatment is challenging. No reference data exists on the implementation of measures for identifying clinical deterioration for adult patients with acute respiratory infections in Latin America.

### STUDY AIM

The aim was to increase the adoption by 50% of evidence-based interventions for early identification of clinical deterioration in adult patients with confirmed or suspected respiratory COVID-19 at the primary level of care after 36 weeks of implementation.

## METHODS

### Study design

We developed a mixed method evaluated Quality Improvement Collaborative (QIC), enrolling nine primary health care centres (PHCC) under an uncontrolled interrupted time-series design with a baseline period (BP) of 12 weeks and an implementation period (IP) of 36 weeks. The project was designed to apply the Breakthrough Innovative Series for the QIC model of the Institute for Healthcare Improvement (IHI). Formal QICs involve healthcare teams from different centres to improve performance on a specific topic by collecting data and testing ideas with plan-do-study-act (PDSA) cycles supported by coaching and learning sessions [7–10].

### Context

We included PHCC of the public sector, which cares for patients with COVID-19 with more than 1,800 visits per month (not expanded due to the pandemic). Based on the eligibility criteria, 9 PHCCs were selected to participate after presenting the project, the expected tasks, and the resources required.

Healthcare workers (HCWs) in the PHCC were the targets of the intervention. Primary and secondary outcomes were measured in patients and HCWs of the participating PHCC. The inclusion criteria for patients were age >18 years and consultation in the participating PHCC for acute respiratory symptoms related to a suspected SARS-CoV 2 infection.

### Intervention

A conceptual framework was established using a driver diagram based on evidence and expert consultation for quality improvement, primary care, and respiratory disease (Supplement, Figure 1). Quality improvement tools were used to develop the framework, including prioritisation matrices and block diagrams.

We implemented a complex toolkit based on several components (Supplement, Table 1).

1. The implementation strategies consisted of selecting a team in each PHCC: an implementation facilitator, data collector, and local study coordinator [11]. The local teams were trained in 1) Audit and feedback to assess the evolution of the package usage rate from study progress reports to establish adoption improvement interventions through PDSA cycles, 2) Site-specific clinical guidelines (CGs) focused on the most frequent circumstances associated with respiratory syndromes in adults[12], 3) Reminders for HCWs, training, and peer coaching[13], 4) Patient and family education about clinical deterioration signs.
2. The intervention comprised four components: 1) A screening tool based on the PRIEST program to triage adult patients with acute respiratory symptoms, based on three colours: green for patients without risk factors and NEWS2 <3, yellow for patients with risk factors and NEWS2 3 or 4, and red for patients with NEWS2≥5, respiratory rate >20/min, unstable comorbidities, or oxygen needs. [14,15], 2) National Early Warning Score 2 (NEWS2) was used periodically to assess the progression of clinical deterioration[16], 3) Portable pulse oximeters were selectively provided to patients with risk factors and NEWS2 3 or 4 for self-monitoring, 4) The provincial telemedicine system was used for patient follow-up between days 5 and 10 after the first consultation to evaluate the evolution and the perception of the received attention in the PHCC[17–20].
3. The intervention was delivered in two presential learning sessions (week 12 and 28). Each learning session constituted the analysis phase of the improvement cycle by discussing the results, lessons learned, interventions’ applicability, and work plan modifications. During the action periods (time between learning sessions), the PHHC’s teams tested and executed the changes in their centres and collected data to measure the impact of the changes, sharing them virtually in monthly virtual sessions with other teams.
4. Quality improvement training was delivered one month prior to the first learning session, virtually through a 10-hour course to provide more tools for system redesign, and its effects were shared in learning sessions to build a community of practice that we left as a lasting result of this project. The course was delivered through four recorded virtual modules and two synchronous virtual sessions, covering the theory and model of improvement, driver diagrams, PDSA cycles, data analysis, and the psychology of change.

### Study of the intervention

Data were extracted from medical records (appointment system, admission records, and follow-up telephone calls between 5-10^th^ day). Run charts illustrating primary outcomes were shared biweekly with teams to monitor progress, and each team reported on the development of improvement opportunities using a standardised report specifically designed for this purpose (Google Classroom™). Control charts were used when enough data was regarded. Audits were conducted in virtually all units every week, and continuous communication was maintained between site coordinators and data collectors via telephone, email, and WhatsApp™.

Semi-structured interviews were conducted with healthcare professionals involved in the intervention implementation at the end of the IP. Interviews included professionals from six participating centres (at least two participants per centre), representing various roles in the study (site coordinators, facilitators, and data collectors) and from different professions (medical, pharmacy, and nursing). Additionally, the study personnel responsible for the telephone follow-up of the enrolled patients were interviewed. A guide was developed for interviews covering topics such as perception of the implementation degree, acceptability and utility of the intervention package and implementation strategies, and barriers and facilitators of study implementation, among other aspects. The interviews were conducted remotely using Zoom. Two qualitative research-experienced investigators who were not involved in the implementation conducted audio-recorded interviews with the participants’ consent.

### Measures

The resulting measure in all enrolled patients was recorded up to the tenth day of follow-up using a risk assessment flowchart based on current local recommendations. The bundle comprised triage and NEWS2 in all suspected or confirmed cases of COVID-19, a portable oximeter as appropriate by the risk category, and 4) a follow-up call.

We reported unexpected events during hospital emergency department visits, such as the occurrence of events such as death and unplanned use of critical interventions (use of vasoactive drugs, noninvasive ventilation, or endotracheal intubation within one hour of arrival at the hospital emergency department) in the reference hospitals. In addition, hospitalisations of the patients enrolled in the study were measured by the frequency of patients admitted to secondary or tertiary-level hospitals during the follow-up period.

As a balancing measure, patients reported their experience with care through a brief survey based on the People Voice Survey of the QuEST network – questnetwork.org delivered by phone calls[21].

Other adjustment variables such as age, sex, clinical status, associated conditions (Charlson Comorbidity Index), health coverage, and educational level were recorded.

### Statistical analysis

Absolute and relative frequencies were reported for categorical variables, whereas means and standard deviations were reported for continuous variables. The chi-square test and t-test were used as appropriate. We used statistical process control to analyse the resulting measures using a p-control chart[22]. To evaluate the effect of the intervention over time, we conducted an interrupted time series (ITS) analysis using a Generalized Least Squares (GLS) model with restricted maximum likelihood estimation. Five different outcomes were considered when implementing each of the four elements and the entire package. Each result was the biweekly proportion, calculated as the number of consultations in which the component or package was implemented, divided by the total number of consultations. The model included three variables: time (fortnight), intervention time, and post-intervention time. This approach allowed us to test the significance of the pre-intervention slope, immediate post-intervention level change, post-intervention slope, and slope changes. When necessary, we accounted for the correlation structure of the errors due to the clustering of observations over time and heteroscedasticity across the intervention periods. Data were analysed using R version 4.0.2 (The R Foundation) and QI Macros for Mac.

In the case of qualitative data, the interviews were transcribed verbatim, uploaded to Atlas.ti and analyzed using thematic content analysis [23].

We used the revised Standards for Quality Improvement Reporting Excellence Guidelines (SQUIRE 2.0) to report this study[24].

### Ethics

The institutional review board approved the study protocol and the informative consent for interviews with users and healthcare workers (Institutional Review Board, Mendoza Government, Ministry of Health, Social Development and Sport, Directorate of Research Science and Technology, Mendoza, Argentina - Disposition number 121/2022 – November 1^st^ 2022).

## RESULTS

### Process measures

Two of the six teams in which the intervention was conducted used all four primary drivers. The other four teams used three primary drivers: improving clinical processes, strengthening staff training and teamwork, and data-driven management. All centres used the primary drivers for improving clinical processes and data-driven management. All teams implemented PDSA cycles to achieve the adoption of triage use for patients presenting with acute respiratory infection and conducted ideas on adopting the use of the NEWS2 score. Only three of the six teams implemented PDSA cycles related to the distribution of oximeters.

### Outcome measures

Data from 877 patients were gathered, 356 in the BP and 477 in the IP. There were no differences between the study groups in age, sex, education level, Charlson score, and referred diagnosis of acute respiratory deterioration (Table 1). During the BP, there were fewer severely obese patients (BP 33.3% vs. IP 48.7%, p=0.047). Eight hundred sixty-two medical consults were reported, 367 in the BP and 495 in the IP. More COVID-19-confirmed diagnoses were observed in the IP group (1.9% vs 15%, p<0.001). More patients required a second consultation in the IP (5.7% vs 10%, p=0.021) (Supplement, Table 3).

**Table 1.** Patient characteristics between study phases.

| Characteristic | Basal (N = 356) | Intervention (N = 477) | p-value* |
| --- | --- | --- | --- |
|  | n/N (%) | n/N (%) |  |
| **Gender** |  |  | 0.791 |
| Male | 115/356 (32.3%) | 152/477 (31.9%) |  |
| Female | 240/356 (67.4%) | 324/477 (67.9%) |  |
| Other | 1/356 (0.3%) | 0/477 (0.0%) |  |
| No respond | 0/356 (0.0%) | 1/477 (0.2%) |  |
| **Age in years\*** | 37.0 (28.0, 49.0) | 37.0 (28.0, 48.0) | 0.828 |
| **Education level** |  |  | 0.519 |
| Never attended | 4/356 (1.1%) | 2/477 (0.4%) |  |
| Incomplete primary | 41/356 (11.5%) | 49/477 (10.3%) |  |
| Complete primary | 80/356 (22.5%) | 90/477 (18.9%) |  |
| Incomplete secondary | 96/356 (27.0%) | 146/477 (30.6%) |  |
| Complete secondary | 97/356 (27.2%) | 133/477 (27.9%) |  |
| Tertiary or University | 38/356 (10.7%) | 57/477 (11.9%) |  |
| **Myocardial infarction** | 1/355 (0.3%) | 0/477 (0.0%) | 0.427 |
| **Congestive heart failure** | 2/355 (0.6%) | 1/477 (0.2%) | 0.579 |
| **Peripheral vascular disease** | 1/355 (0.3%) | 11/477 (2.3%) | 0.015 |
| **Cerebrovascular disease** | 1/355 (0.3%) | 1/477 (0.2%) | >0.999 |
| **Dementia** | 0/355 (0.0%) | 0/477 (0.0%) | >0.999 |
| **Chronic Lung Disease** | 13/355 (3.7%) | 22/477 (4.6%) | 0.500 |
| **Connective tissue pathology** | 0/355 (0.0%) | 1/477 (0.2%) | >0.999 |
| **Ulcer disease** | 0/355 (0.0%) | 0/477 (0.0%) | >0.999 |
| **Mild liver pathology** | 0/355 (0.0%) | 2/477 (0.4%) | 0.510 |
| **Moderate to severe liver pathology** | 0/355 (0.0%) | 1/477 (0.2%) | >0.999 |
| **Diabetes** | 25/355 (7.0%) | 40/477 (8.4%) | 0.475 |
| **Diabetes with organic damage** | 1/355 (0.3%) | 2/477 (0.4%) | >0.999 |
| **Hemiplegia** | 1/355 (0.3%) | 0/477 (0.0%) | 0.427 |
| **Moderate or severe kidney pathology** | 2/355 (0.6%) | 2/477 (0.4%) | >0.999 |
| **Neoplasms** | 0/355 (0.0%) | 1/477 (0.2%) | >0.999 |
| **Leukemia** | 0/355 (0.0%) | 0/477 (0.0%) | >0.999 |
| **Malignant lymphomas** | 1/355 (0.3%) | 1/477 (0.2%) | >0.999 |
| **Solid metastasis** | 0/355 (0.0%) | 0/477 (0.0%) | >0.999 |
| **AIDS** | 2/355 (0.6%) | 1/477 (0.2%) | 0.579 |
| **None of the above** | 307/355 (86.5%) | 403/477 (84.5%) | 0.422 |
| **Charlson ICM Score\*** | 0.00 (0.00, 1.00) | 0.00 (0.00, 1.00) | 0.937 |
| **Estimated survival at 10 years (Charlson score) \*** | 0.98 (0.96, 0.98) | 0.98 (0.96, 0.98) | 0.937 |
| **Obesity** | 112/246 (45.5%) | 161/412 (39.1%) | 0.104 |
| **Degree of obesity€** |  |  | 0.047 |
| Grade I or moderate (BMI 30 – 34.9) | 46/96 (47.9%) | 39/119 (32.8%) |  |
| Grade II or severe (BMI 35-39.9) | 32/96 (33.3%) | 58/119 (48.7%) |  |
| Grade III or morbid (BMI ≥ 40) | 18/96 (18.8%) | 22/119 (18.5%) |  |
| **Pregnancy** | 6/230 (2.5%) | 3/322 (0.9%) | 0.174 |

\*Mean (SD), €Denominator: Participants with Obesity

**Table 1.** Interrupted time series analysis for the complete bundle.

|  | Complete Bundle |  | Triage use |  | NEWS2 score use |  | Follow-up call use |  |
| --- | --- | --- | --- | --- | --- | --- | --- | --- |
|  | Coefficient<br>(95% CI) | P-value | Coefficient<br>(95% CI) | P-value | Coefficient<br>(95% CI) | P-value | Coefficient<br>(95% CI) | P-value |
| **Baseline level of the outcome** | 0.00<br>(0.00;0.23) | 1.00 | 0.01<br>(0.00;0.02) | 0.88 | 0.17<br>(0.10;0.24) | <0.01 | 0.72<br>(0.59;0.85) | <0.01 |
| **Slope prior to the intervention** | -0.00<br>(-0.06;0.06) | 1.00 | 0.01<br>(-0.01;0.01) | 0.79 | -0.03<br>(-0.05;-0.02) | 0.01 | -0.05<br>(-0.09;-0.02) | <0.01 |
| **Change in level immediately after the intervention** | 0.20<br>(-0.01;0.41) | 0.08 | 0.32<br>(0.21;0.43) | <0.01 | 0.64<br>(0.55;0.73) | <0.01 | 0.23<br>(0.08;0.37) | <0.01 |
| **Change in the slope from pre to post intervention** | 0.01<br>(-0.05;0.06) | 0.84 | 0.03<br>(0.02;0.04) | <0.01 | 0.06<br>(0.04;0.08) | <0.01 | 0.04<br>(0.01;0.07) | 0.04 |
| **Slope post to the intervention** | 0.01<br>(-0.01;0.02) | 0.30 | 0.03<br>(0.02;0.04) | <0.01 | 0.03<br>(0.02;0.032) | <0.01 | -0.02<br>(-0.03;-0.01) | <0.01 |

\*A Generalized Least Squares model adjusting for heteroscedasticity across intervention periods was used considering an autocorrelation ARIMA(3,0).

All four components of the bundle were implemented in 0% and 24.8% of the patients in the BP and IP groups, respectively (Figure 2). In the interrupted time-series analysis, there were no significant changes in the level immediately after the implementation or in the slope from BP to IP (Table 2). Upon evaluating the individual components of the bundle, we discovered enhancements in the utilisation of triage (0.5% vs 57%), the application of the NEWS2 score (4.1% vs 80.0%), and the utilisation of oximeters when applicable (0% vs 48%). We observed a slight decrease in follow-up calls, as shown in (Figure 3). We observed an improvement with significant changes in the slope and level immediately after the implementation and changes in the slope from pre-to post-implementation using triage and the NEWS2 scale. A decrease in follow-up calls was shown, with significant changes in the slope and level immediately after the intervention, as well as changes in the slope from pre-to post-intervention (Table 2).

**Figure 1.**
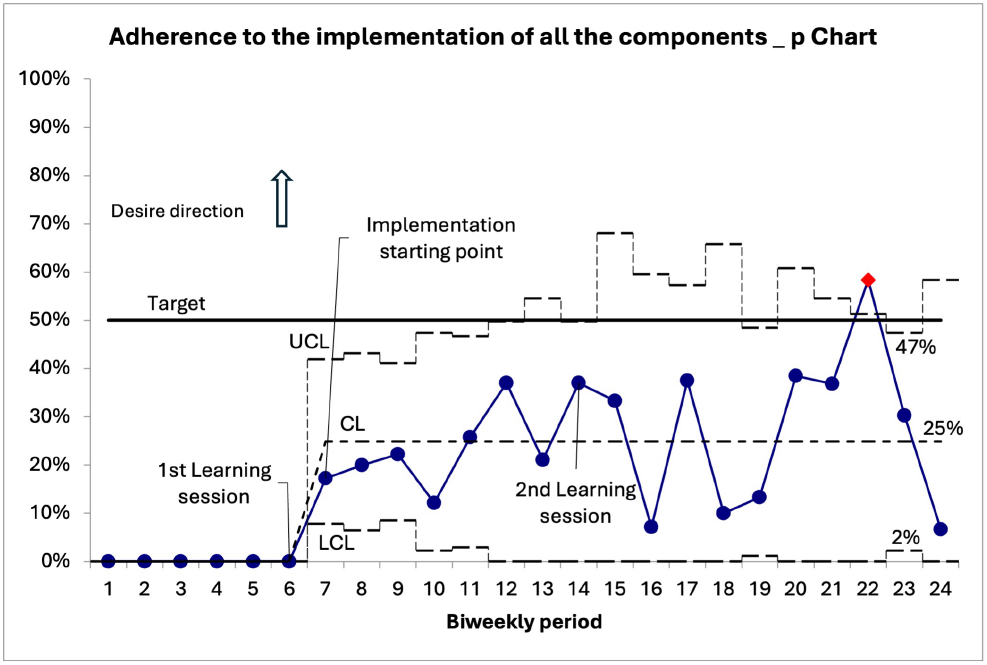
P-Chart of the adherence of the four components of the implementation bundle (triage, NEWS2 score, oximeter use and follow-up calls) during the study period (red line, special variation; blue line, expected or common variation).

**Figure 2.**
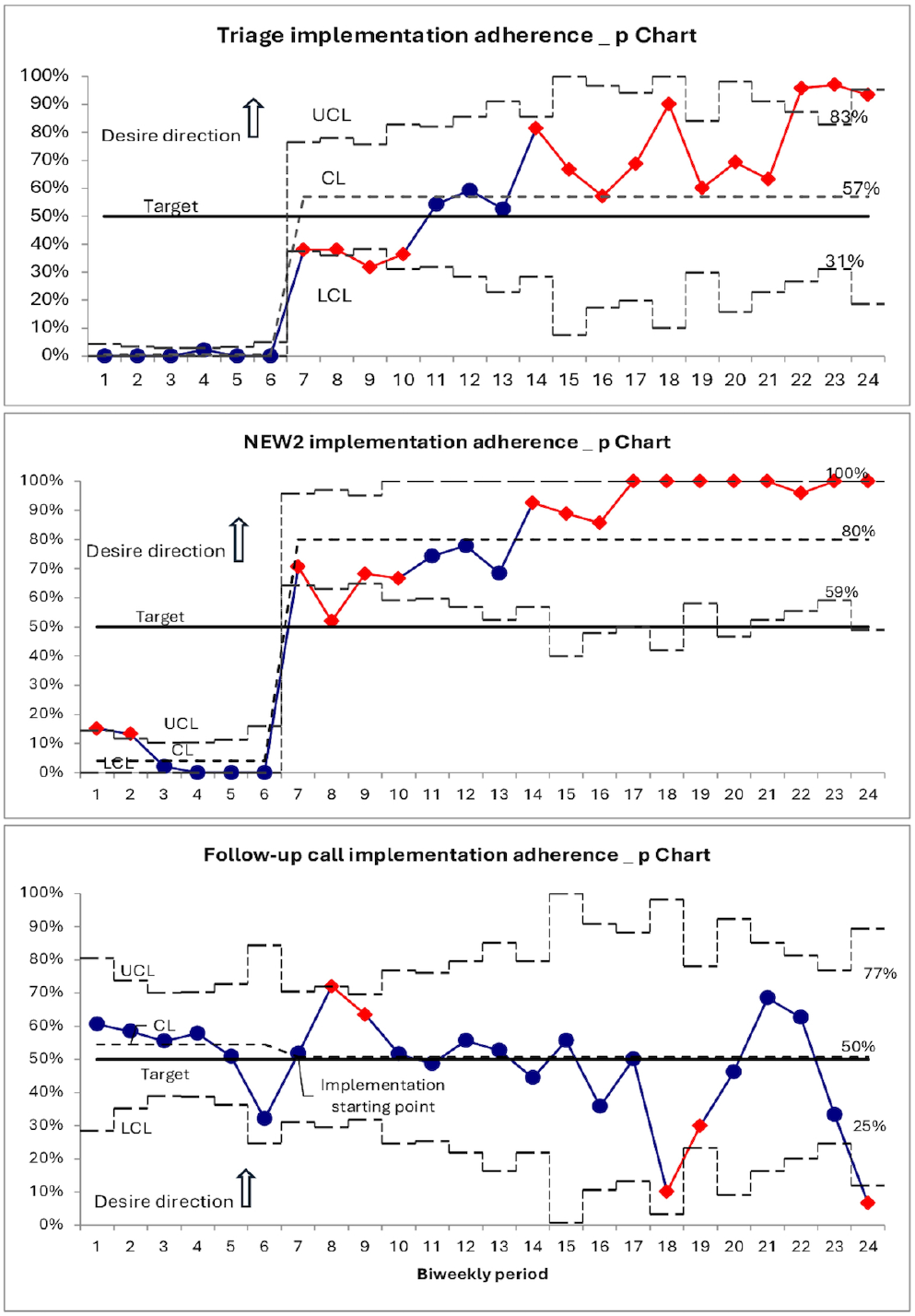
P-charts of the implementation adherence of the different components of the bundle A) Triage B) NEWS 2 score C) Follow-up call. In red, special variation; in blue, expected or common variation.

**Table 2.** People voice survey concerning care experience.

| Question | Basal<br>(N = 195) | Intervention<br>(N = 245) | p-value* |
| --- | --- | --- | --- |
|  | n (%) | N (%) |  |
| **How would you rate the quality of care in general?** |  |  |  |
| Positive | 128 (66%) | 185 (76%) | 0.023 |
| **How would you rate the knowledge and ability of the health personnel who treated you?** |  |  |  |
| Positive | 147 (75%) | 202 (82%) | 0.069 |
| **How would you rate the medical equipment and items available, such as access to laboratory tests?** |  |  |  |
| Positive | 110 (56%) | 145 (59%) | 0.558 |
| **How would you rate the respect shown to you by the health personnel?** |  |  |  |
| Positive | 155 (79%) | 204 (83%) | 0.310 |
| **How would you rate the knowledge of health personnel about your previous consults or examinations?** |  |  |  |
| Positive | 133 (68%) | 158 (64%) | 0.413 |
| **How would you rate the staff's ability to explain things to you in a way you could understand?** |  |  |  |
| Positive | 145 (74%) | 199 (81%) | 0.083 |
| **How would you rate the ability of health personnel to involve you in decisions about treatment?** |  |  |  |
| Positive | 139 (71%) | 184 (75%) | 0.368 |
| **How would you rate the time the staff dedicated to you?** |  |  |  |
| Positive | 137 (70%) | 199 (81%) | 0.007 |
| **How would you rate the time you had to wait before the consult?** |  |  |  |
| Positive | 92 (47%) | 165 (67%) | <0.001 |
| **How would you rate the kindness and help that the rest of the staff there gave you?** |  |  |  |
| Positive | 139 (71%) | 185 (76%) | 0.317 |
| **How would you rate the information on alarm guidelines to take into account in relation to the disease for which you consulted?** |  |  |  |
| Positive | 140 (72%) | 179 (73%) | 0.768 |
| **If you have received a pulse oximeter, how would you rate the information received about its use?** |  |  |  |
| Positive | 8 (4.1%) | 24 (9.8%) | 0.022 |
| *Pearson's Chi-squared test was used. |  |  |  |

The distribution of triage colour in the IP group was 87.9% (248) green, 11% (31) yellow, and 1.1% (3) red. In the BP group, only two patients received triage; both were classified as yellow. There were no differences in patient hospitalisations between the study groups (BP 0% vs IP 1.6%, RR 7.17 (95%CI 0.39;132.39), p= 0.185).

### Balancing measure

Patients rated the quality of care as positive in 66% of the cases in the BP and 76% in the IP group (p=0.023). Furthermore, the percentage of time the staff dedicated was improved in the IP group (70% vs. 81%, p=0.007). Moreover, the IP group rated waiting time before consultation more positively (47% vs. 67%, p<0.001). The remaining questions from the survey are presented in Table 3.

### Qualitative results

In the qualitative component of the study, 23 healthcare professionals provided insights into the implementation process (Supplement, Table 4). The participants highlighted the benefits of the intervention package, particularly its role in improving care organisation, standardising procedures, and enhancing the management of respiratory deterioration. Patient recruitment faced barriers, including restrictive inclusion criteria and limited collaboration with referral services. Challenges in adoption were noted, as implementation often depended on a few active team members, with limited engagement from the broader healthcare staff and a lack of endorsement from local health authorities and centre leadership. Despite these challenges, the involvement of non-medical staff, especially nurses, was identified as a facilitator for successful implementation. Detailed qualitative findings will be reported separately.

## DISCUSSION

The primary objective of this study was to accomplish a complete bundle of four components when appropriate. In most cases, the centres could fulfil three components: triage use, NEWS2 score use, and the use of a portable oximeter when it has been provided as appropriate by the risk category. However, the follow-up calls showed a decrease in compliance during implementation. A non-significant improvement was observed regarding the complete use of the bundle, considering that 50% of use was not performed. Some progress was made in the home utilisation of pulse oximeters, although this development did not reach our target of 50%. Moreover, patients reported more favourable care experiences during the implementation period.

Using the triage algorithm assessment, respiratory symptoms were graded and placed in the context of patient risk factors. Only three patients were rated red in the triage and were hospitalised. No patient experienced deterioration, which prevented us from concluding the usefulness of the tools in identifying deterioration. All teams implemented PDSA cycles to achieve the adoption of triage use for patients presenting with acute respiratory infection and conducted ideas on adopting the use of the NEWS2 score. These elements were most related to usual practices. However, there are valid clinical reasons why deteriorated patients may not have been transported to the hospital. Implementing any triage tools could minimise the likelihood of false-negative triage and the non-conveyance of a patient who subsequently experiences a decline in their condition [25].

Home pulse oximetry can help to monitor breathlessness. These were recommended for post-acute COVID-19 management in primary care settings[26]. Only 31 patients (11%) presented with yellow during the IP triage and 2 in the BP triage. Nearly 50% of the yellow-triaged patients received an oximeter to monitor pulse oximetry at home. None of the patients required further hospitalisation. Using home pulse oximetry could spare hospital resources for individuals who could greatly benefit from an escalation of care[27]. Unfortunately, it was impossible to determine unequivocal proof of the impact of pulse oximetry on the specified results.

Implementing follow-up calls was the most challenging aspect of this study. Despite our best efforts, the local telehealth system was never established because of a lack of budget, ultimately leading to our efforts’ discontinuation by the end of the study.

Implementing the intervention improved patient satisfaction with the quality of care provided. Specifically, the patients rated the time dedicated to them and the waiting time before their consultation as the two most positive aspects. The standardisation of care and involvement of non-medical personnel contributed to these positive ratings.

This research constitutes one of the initial studies conducted in a Latin American country to assess the implementation of a bundle to identify respiratory deterioration in patients with COVID-19 or other respiratory infections in PHHCs. We used a strength measurement strategy to evaluate the implementation and people voice surveys as a balancing measure. Moreover, we applied a QIC based on the Breakthrough Innovation series of the IHI, which has demonstrated favourable outcomes in our region[28,29]. The utilisation of the triage algorithm founded on NEWS2 in primary care settings has garnered extensive approval, thereby enabling the expansion of interventions to additional locations within the region.

### Limitations

Three centres abandoned the study. One was closed for building renovation work. Another team collected data but had to quit because of staff restructuring. The last had problems forming a team at the beginning of the study. All centres had staff turnover due to the country’s economic problems. These changes forced us to provide repeated training to the data collectors and intervention facilitators. Establishing a local telehealth system presented significant challenges. To address this, we enlisted the support of healthcare professionals from a single facility and medical career students who aided in conducting follow-up calls. Unfortunately, our capacity to perform follow-up calls was restricted to the fifth to tenth days post-intervention, and we could only make a single call per patient.

## CONCLUSIONS

We implemented a triage algorithm based on the NEWS2 score to identify respiratory deterioration in adult patients in primary care through a QIC. This intervention improved the patient’s perceptions of their quality of care. In addition, we provided home pulse oximetry to patients who were more likely to develop deterioration. However, we were unable to determine the effectiveness of home pulse oximetry. It is worth noting that implementing follow-up calls required a significant commitment from local health authorities.

## Data Availability

All data produced in the present study are available upon reasonable request to the authors

https://osf.io/dnpsq/?view_only=9b336ab2a9f9496f99c9ccccc2c0aeb6

