## Supplement for "Timely Identification of deteriorating Patients from acute respiratory infections at the primary care level in the COVID-19 Era: quality improvement collaborative"

#### Content table

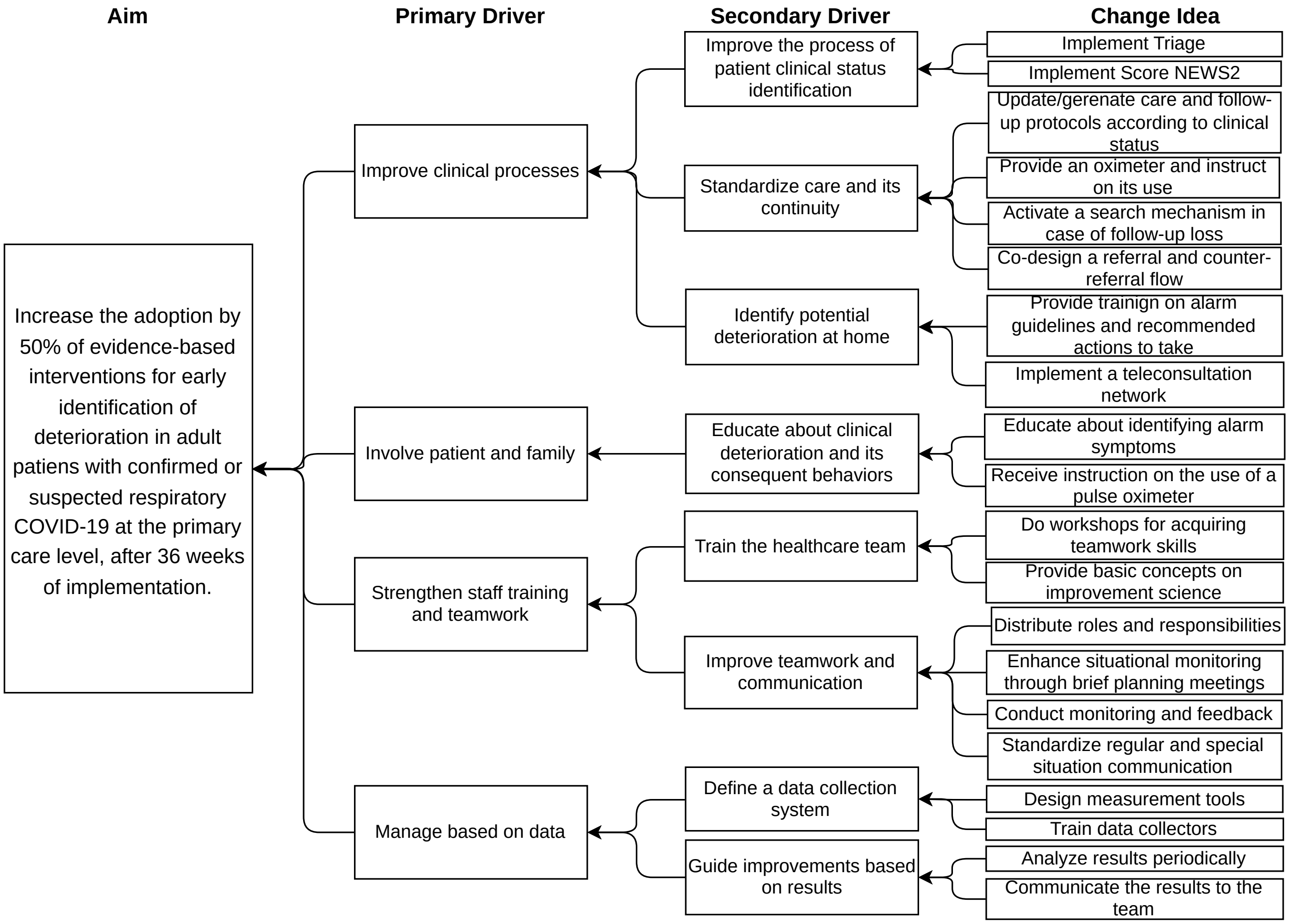

*Figure 1. Driver diagram explaining the theory of change in study intervention.*

*Table 1. The TIDieR (Template for Intervention Description and Replication) Checklist*

| **BRIEF NAME** |
|---|
| Timely detection of deterioration in patients with acute respiratory infection in primary care. |
| **WHY** |
| At the individual patient with acute respiratory infection: to ensure standardized and evidence-based care for these patients to conduct proper assessment and timely detection of clinical deterioration, to take appropriate actions to prevent unexpected outcomes.<br>At the health system level: to contribute to a model of care that is prompt, efficient, and safe at the primary care level, strengthening capacities to manage peaks in incidence of acute respiratory infections caused by SARS-CoV-2 and other pathogens. This aims to prevent overburdening of secondary and tertiary levels of care. |
| **WHAT** |
| **The key components included:**<br>- **Conceptual framework:** A key driver diagram (KDD) was created to establish the conceptual framework based on evidence and discussions with experts in quality improvement and primary healthcare providers.<br>- **Quality improvement tools:** prioritization matrix, block diagrams, Plan-Do-Study-Act (PDSA) cycles.<br>- **Conformation of a local improvement team in each center**<br>- **Quality improvement course**<br>- **In-person learning sessions**<br>- **Monthly virtual sessions:** to share knowledge and experience in a collaborative manner.<br>- **Coaching support:** in one-to-one biweekly meetings with primary health care centers.<br>- **Measurement strategy**<br>- **Virtual platform:** repository of materials including templates for recording improvement cycles (Google Classroom™).<br>- **Training in concepts of institutionalization and sustainability of improvements.**<br>- **Package of specific tools** for the evidence-based evaluation and clinical monitoring of patients with acute respiratory infection.<br> ● **Triage of adult patients with acute respiratory symptoms:** Use of the screening tool based on the PRIEST program (Bentivegna, Hulme, and Ebell, 2021; Marincowitz et al., 2022). It was based on three colors: green for patients without risk factors and NEWS2 <3, yellow for patients with risk factors and NEWS2 3 or 4, and red for patients with NEWS2 ≥5, respiratory rate >20/min, unstable comorbidities, or oxygen needs.<br> ● **Use of a deterioration scale for clinical monitoring:** To assess the progression of deterioration in patients at higher risk, the National Early Warning Score 2 (NEWS2) was used periodically. This scale combines six physiological variables, whose individual scores are summed to obtain an aggregate score, and higher scores are associated with greater severity of the disease. (Royal College of Physicians, 2017; Zaman et al., 2021).<br> ● **Supply and use of portable oximeters**: Saturation meters was selectively provided to patients at higher risk for self-monitoring. The remote patient monitoring seems to be effective in helping triage patients, thus allowing health services to be prioritized for the people who need them most. Home oximetry could help prevent the unnecessary use of emergency services and identify deteriorating patients in a timely manner, avoiding treatment delays and prolonged hospital admissions. (Mohammadzadeh and Safdari, 2014; Noah et al., 2018; Torjesen, 2020; Alboksmaty et al., 2022)<br> ● **Follow-up based on clinical criteria**. |

- **Telephone contact:** to assess the care experience users in the primary care setting and the progression of the illness.

**WHO PROVIDED**

The study intervention required the coordinating team to have an expert-level understanding of quality improvement and implementation, the local improvement team to possess basic-level knowledge with supervision by coaching experts in quality improvement. The coordinating team was formed by 8 people with specialties in quality improvement, improvement coaching, primary care, emergency care and organizational leadership. PHCC teams were formed by physicians, nurses, clerks and pharmacist.

**HOW**

**Quality improvement course:** consisted of four asynchronous virtual modules and two synchronous virtual sessions. It covered a wide range of topics, including the theory of improvement, the model of improvement, KDD, PDSA cycles, data analysis, and the psychology of change. The course had a total duration of 40 hours.

**In-person learning sessions** with training and coaching for improvement teams. Two in-person learning sessions were conducted in which staff were trained in quality improvement concepts and the psychology of change. Practical workshops were also held to work on tools for implementing improvement cycles (PSDA).

**Virtual sessions for consolidating improvement development**. These monthly learning activities lasted 60 minutes and involved all centers and the coordinating team. The purpose of these activities was to promote collaborative learning and networking among all the centers and the study's coordinating team, focusing on biweekly data and ongoing efforts. During these sessions, each participating center would present an improvement cycle to foster collective learning and exchange ideas on challenges and strategies for implementing changes. Additionally, updates on global measurements were shared.

**Support for centers by a coach.** They met virtually every two weeks with each center, assisting the improvement team in the development and monitoring of improvement cycles, and jointly analyzing the data collected at that center to guide their interventions based on that data.

**Implementation deployment (change ideas):**

- Each site formed teams with local facilitators, with at least one member of the healthcare team in the role of executive implementation leader, responsible for planning, dissemination, and development.
- The overall coordination of the study was handled by specialists in healthcare quality and coaching for improvement.
- Multidisciplinary teams composed of nurses, doctors, and clerks from HPCCs participated in the co-design of the intervention at each site. According to the reality and needs of their center, each team selected change ideas proposed in the KDD and conducted PDSA improvement cycles, setting goals and tasks for each idea with designated responsibilities and timelines to achieve the objective.
- Follow-up calls were made by a coach. The objectives of the calls were to provide comprehensive support for implementation, analyze measurement results, and identify challenges in the ongoing PDSA cycles to agree on the next steps.
- Each center used Google Classroom™ to record the PDSA cycles.

**Data for improvement:** we reported ongoing global data and for each site in a biweekly manner. The data for each outcome were consolidated in run-charts.

**WHERE**

Interactions between the coordinating group, the sites, and among the sites themselves were conducted virtually. Coaching sessions were conducted remotely via Zoom®. Learning sessions were conducted in-person in Mendoza with all the participant PHCC teams

**WHEN AND HOW MUCH**

- The quality improvement implementation course comprises four modules, each lasting 5 educational hours, and includes two synchronous virtual sessions, each spanning 90 minutes. Reading and learning materials are accessible on the virtual campus, providing permanent access for participants.
- Virtual sessions occur monthly, with each session lasting 60 minutes, commencing from week 12 and continuing until week 52 with the aim of assessing sustainability.
- Biweekly coaching sessions were provided, with durations ranging from 30 to 60 minutes, beginning at week 12 and continuing until week 48.
- Additionally, two extended in-person learning sessions was conducted, each lasting 240 minutes.

**TAILORING**

Coaching sessions were customized to suit the specific requirements, considering the site's level of expertise in implementing quality improvements.
The improvement suggestions were not rigidly defined, permitting each center to adjust and apply the ideas in alignment with their own capabilities and starting point.

**MODIFICATIONS**

*None*

**HOW WELL**

**Participation**: Three of the health centers did not collect data or implement the intervention. One was closed for building renovation work. Another team collected data but had to quit because of staff restructuring. The last had problems forming a team at the beginning of the study.
**Theory of change:** The KDD featured four primary drivers, 8 secondary drivers, and 20 change ideas. It was anticipated that by the study's conclusion, all sites would have implemented at least one test or change idea for each primary driver.

- **Change ideas implementation**: among the 6 teams where the intervention was conducted, 2 used all 4 primary drivers. The other 4 teams used 3 of the 4 primary drivers: improving clinical processes, strengthening staff training and teamwork, and data-driven management. Only the primary drivers of improving clinical processes and data-driven management were used by all the centers that received the intervention.
- **Implementation of triage use**: all teams implemented improvement cycles to achieve the adoption of triage use for patients presenting with acute respiratory infection.
- **Implementation of NEWS2 score use:** 100% of the teams that conducted the intervention worked on adopting the use of the NEWS2 score for patients with acute respiratory infection.
- **Distribution of oximeters:** only 3 of the 6 teams implemented improvement cycles related to the adoption of this element.
- **Data-driven management:** All centers were sent a biweekly report with data corresponding to their center and global data. This data was used to manage the selection of change ideas to work on. Data entry training was conducted in all centers. Additionally, the last 2 monthly exchange sessions focused on sustainability and institutionalization of change, providing possible measurement tools that the centers could adopt.

*Table 2. Patient risk factors*

| Patient Risk Factors |
|---|
| >65 years |
| Diabetes mellitus type 1 or 2, |
| Obesity grade 2 or 3 |
| Chronic cardiovascular disease |
| Chronic kidney disease |
| Chronic respiratory disease |
| Chronic liver disease |
| Patients on the waiting list for solid organ transplants and solid organ transplant recipients |
| People with disabilities, residents of nursing homes, residences, and small homes |
| Oncological and oncohematological patients with a recent diagnosis or “ACTIVE” disease |
| People with active tuberculosis |
| Down syndrome |
| People with autoimmune diseases and/or immunosuppressive, immunomodulatory or biological treatments |
| Pregnant and postpartum people |

*Table 3. Medical consultation characteristics between the study phases*

| Characteristic | Basal (N = 367) | Intervention (N = 495) | p-value* |
|---|---|---|---|
| | n/N (%) | n/N (%) | |
| **COVID-19 situation** | | | <0.001 |
| Suspicious | 66/367 (18%) | 27/495 (5.5%) | |
| Confirmed | 7/367 (1.9%) | 75/495 (15%) | |
| Unsuspicious | 55/367 (15%) | 98/495 (20%) | |
| Not reported | 30/367 (8.2%) | 35/495 (7.1%) | |
| Negative | 209/367 (57%) | 260/495 (53%) | |
| **Referred acute respiratory diagnoses** | | | 0.119 |
| Upper respiratory disease | 186/356 (52%) | 227/477 (48%) | |
| Influenza-like illness | 53/356 (15%) | 58/477 (12%) | |
| Acute Obstructive Bronchial Syndrome | 8/356 (2.2%) | 18/477 (3.8%) | |
| Acute bronchitis | 34/356 (9.6%) | 59/477 (12%) | |
| Pneumonia | 4/356 (1.1%) | 15/477 (3.1%) | |
| Other | 71/356 (20%) | 100/477 (21%) | |
| **Requires a new consultation** | 21/367 (5.7%) | 50/495 (10%) | 0.021 |

*Table 4. Interviewed healthcare workers' characteristics*

| Characteristics | n |
|---|---|
| **Sex** | |
| - **Female** | **15** |
| - **Male** | **8** |
| **Profession** | |
| - **Physician** | **8** |
| - **Nurses** | **4** |
| - **Pharmacists and biochemists** | **3** |
| - **Administrative** | **4** |
| - **Medical students** | **5** |
| **Role in the study** | |
| - **Site coordinators** | **4** |
| - **Facilitators** | **4** |
| - **Data collection** | **9** |
| - **Patient follow-up via phone** | **5** |
| **Total** | **23** |

### Appendix

#### Interview Guide for Healthcare Personnel (English version)

1. Describe your experience working with patients with respiratory symptoms.
2. Considering what happened during the pandemic, what measures or restructuring were adopted to care for patients with respiratory infections during the COVID-19 pandemic?
3. Did your attitude/approach change as the situation progressed?
4. How do patients with symptoms of respiratory infection visit the consult?
    a. What is your perception of the risk to patients with respiratory symptoms?
    b. What is your opinion on the information that patients receive regarding their respiratory symptoms?
5. What tests are currently used to detect infections? In Primary Care, in Secondary Care?
6. What tests are currently used to detect COVID-19? In Primary Care, in Secondary Care?
    a. How reliable are these results?
    b. Who participated in the sample collection process (number of people, training, and background)?
    c. Do you believe that the current tests meet the requirements of a Primary Care setting? In Secondary Care settings
    d. What is the average time from conducting the tests to receiving results?
    e. Who receives the results, and what procedures exist to act on them?
7. What treatment do the patients receive?
    a. Referrals to other levels, other services
    b. Follow-up
8. What challenges or obstacles do you face regarding the provision of assistance?
9. What obstacles do you encounter concerning coordination between different levels, health centres, and programs?

10. In your opinion, what factors contribute to good care in your health centre?
    a. What could promote the use and acceptance of your medical centre?
11. If the demand suddenly changes during normal operations regarding cases of infection, how would you manage it?
12. You have gained considerable experience during the pandemic. Assuming there could be other pandemics in the future, what should be done to ensure that primary care is prepared?
13. Do you think the number of people undergoing testing will increase in the winter? If so, to what extent?
14. In your opinion, where are the issues in decision-making and current testing?
15. I would like you to reflect on seeking help for a lung infection. What do you think would prevent someone from seeking help when they have symptoms indicative of lung infection?
    a. What do you think encourages someone to seek help?
    b. What do you think can overcome these obstacles?
16. Is there anything else you would like to add?
